# Risk of vaccine preventable diseases in UK migrants: a serosurvey and concordance analysis, 2020

**DOI:** 10.1101/2021.05.04.21253031

**Authors:** Mayuri Gogoi, Christopher A. Martin, Martin J. Wiselka, Judi Gardener, Kate Ellis, Valerie Renals, Adam J. Lewszuk, Sally Hargreaves, Manish Pareek

## Abstract

We conducted a serosurvey in 2020, amongst 149 adult migrants living in the United Kingdom, to determine seroprotection rates for measles, varicella zoster, and rubella. Findings suggest a gap in seroprotection against measles (89.3%). Younger migrants and those from Europe and Central Asia may be more susceptible; self-reported vaccine/disease status is a poor predictor of seroprotection. Understanding factors associated with seroprotection among migrants is critical for informing the delivery of SARS-CoV-2 vaccine.

---

In 2018, the United Kingdom (UK) lost its measles elimination status owing to a steep rise in cases (1). Unvaccinated individuals are considered to be particularly vulnerable to these outbreaks. Migrants may represent an under-immunised group and therefore at increased risk of acquiring Vaccine Preventable Diseases (VPDs) (2, 3). However, there is a paucity of data relating to VPD seroprotection rates and demographic predictors of seroprotection against VPDs in migrants living in the United Kingdom (UK). Determining factors associated with a lack of seroprotection (which are likely to be similar to factors associated with a lack of vaccination) in migrants is critical to inform SARS-CoV-2 vaccine delivery in this population. We, therefore, conducted a rapid cross-sectional serosurvey, to establish seroprotection rates and predictors of seroprotection against measles, rubella and varicella in an adult migrant population in the UK and investigated the concordance of self-reported vaccination and disease history with serostatus.

## Setting and Investigation

The study was conducted in Leicester, UK where approximately 34% of the population are foreign-born (4). We included adult migrants (≥16 years) residing in Leicester (those staying <6months were excluded). Recruitment was at four different sites (see supplementary information). Participation was voluntary and written informed consent was obtained from all participants. Ethical approval was received from HRA and Health and Care Research Wales (reference number 19/LO/1846).

Participants completed a questionnaire in English and provided a blood sample which was analysed for the presence of IgG against measles, varicella and rubella at University Hospitals of Leicester NHS Trust (see supplementary information). Demographic information (age, sex, ethnicity, year of UK arrival and immigration status on arrival) as well as self-reported history of infection and vaccination against measles, varicella and rubella were collected in the questionnaire. If year of UK arrival was missing we accessed the electronic health record and used the first contact with health services as a proxy measure.

We summarised the data using median and interquartile range (IQR) for non-parametric continuous variables and count and percentage for categorical variables. We used univariable and multivariable logistic regression to determine predictors of (i) seroprotection against all studied VPDs and (ii) seroprotection against measles. Agreement between self-reported vaccination and disease history and IgG result were assessed using Cohen’s kappa coefficients (K) and positive and negative predictive values (PPV and NPV respectively). Analyses were conducted using Stata (StataCorp LP, Texas, USA, Version 15.1), p values < 0.05 were considered significant.

## Characteristics of study participants

150 participants were recruited. One blood sample was discarded due to a labelling error and excluded from analysis resulting in a final cohort of 149 participants. Table 1 shows a description of the cohort, median age was 38 years and the majority (67.8%) were female. Amongst migrants from Europe and Central Asia, 32/38 (84.2%) were from Eastern Europe.

**Table 1.** Description of cohort stratified by serology result.

|  | Total<br>n=149 | Measles |  | Rubella |  | Varicella |  |
| --- | --- | --- | --- | --- | --- | --- | --- |
|  |  | IgG positive<br>n=133 (89.3%) | IgG negative<br>n=16 (10.7%) | IgG positive<br>n=138 (92.6%) | IgG negative<br>n=11 (7.4%) | IgG positive<br>n=146 (98.0%) | IgG negative<br>n=3 (2%) |
| **Age, years (med IQR)** | 38 (31 – 48) | 39 (32 – 49) | 33.5 (22.5 – 39) | 38 (31 – 48) | 37 (26 – 43) | 38.5 (31 – 48) | 21 (20 – 33) |
| **Sex, n (%)** |  |  |  |  |  |  |  |
| Male | 48 (32.2%) | 46 (95.8%) | 2 (4.2%) | 44 (91.7%) | 4 (8.3%) | 47 (97.9%) | 1 (2.1%) |
| Female | 101 (67.8%) | 87 (86.1%) | 14 (13.9%) | 94 (93.1%) | 7 (6.9%) | 99 (98.0%) | 2 (2.0%) |
| **Region of origin, n (%)** |  |  |  |  |  |  |  |
| Europe & Central Asia | 38 (25.5%) | 28 (73.7%) | 10 (26.3%) | 37 (97.4%) | 1 (2.6%) | 38 (100.0%) | 0 (0.0%) |
| Africa & Middle East | 35 (23.5%) | 34 (97.1%) | 1 (2.9%) | 33 (94.3%) | 2 (5.7%) | 35 (100.0%) | 0 (0.0%) |
| South Asia, East Asia & Pacific | 76 (51.0%) | 71 (93.4%) | 5 (6.6%) | 68 (89.5%) | 8 (10.5%) | 73 (96.1%) | 3 (4.0%) |
| **Immigration status on arrival, n (%)** |  |  |  |  |  |  |  |
| UK national | 4 (2.7%) | 4 (100.0%) | 0 (0.0%) | 4 (100.0%) | 0 (0.0%) | 4 (100.0%) | 0 (0.0%) |
| Indefinite or discretionary leave to remain | 7 (4.7%) | 6 (85.7%) | 1 (14.3%) | 7 (100.0%) | 0 (0.0%) | 6 (85.7%) | 1 (14.3%) |
| Study or work visa | 7 (4.7%) | 7 (100.0%) | 0 (0.0%) | 5 (71.4%) | 2 (28.6%) | 7 (100.0%) | 0 (0.0%) |
| Husband / wife sponsorship | 38 (25.5%) | 36 (94.7%) | 2 (5.3%) | 35 (92.1%) | 3 (7.9%) | 37 (97.4%) | 1 (2.6%) |
| EEA national – working / self-supporting / on welfare / studying | 49 (32.9%) | 39 (79.6%) | 10 (20.4%) | 46 (93.9%) | 3 (6.1%) | 49 (100.0%) | 0 (0.0%) |
| Refugee / asylum seeker / humanitarian protection | 16 (10.7%) | 15 (93.8%) | 1 (6.3%) | 16 (100.0%) | 0 (0.0%) | 16 (100%) | 0 (0.0%) |
| Don't know / prefer not to say / other | 28 (18.8%) | 26 (92.9%) | 2 (7.1%) | 25 (89.3%) | 3 (10.7%) | 27 (96.4%) | 1 (3.6%) |

## Rate and predictors of seroprotection

Seroprotection rates were: varicella zoster 98%, rubella 92% and measles 89.3% [with only measles being below its herd immunity threshold (HIT) of 95% (5)] (Table 1). Seroprotection rates for measles were lower amongst migrants from Europe and Central Asia compared to those from Africa and the Middle East (73.7% vs 97.1% [chi-square p=0.005]) or South Asia, East Asia and the Pacific (73.7% vs 93.4% [chi-square p=0.003]).

Adjusted and unadjusted analysis of factors associated with seroprotection against all studied VPDs is shown in Supplementary Table 2. Older migrants were more likely to have seroprotection (aOR 1.07 95%CI 1.01 – 1.14 for each year increase in age).

On adjusted analysis, compared to those from Europe and Central Asia, migrants from Africa and the Middle East (aOR 18.32 95%CI 1.63–206.09) and South Asia, East Asia and Pacific regions (aOR 12.75 95%CI 2.25–72.27) were significantly more likely to be seroprotected against measles (Table 2).

**Table 2.** Unadjusted and adjusted analysis of factors associated with seroprotection against measles.

| Variable | n seroprotected/total (%)<br>133/149 (89.3%) | Unadjusted OR (95% CI) | p value | Adjusted OR (95% CI) | p value |
| --- | --- | --- | --- | --- | --- |
| **Age** | - | 1.07 (1.01 – 1.13) | 0.01 | 1.23 (1.09 – 1.38) | 0.001 |
| **Sex** |  |  |  |  |  |
| Male | 46 / 48 (95.8%) | - | - | - | - |
| Female | 87 / 101 (86.1%) | 0.27 (0.06 – 1.24) | 0.09 | 0.23 (0.04 – 1.35) | 0.10 |
| **Region of origin** |  |  |  |  |  |
| Europe & Central Asia | 28 / 38 (73.7%) | - | - | - | - |
| Africa & Middle East | 34 / 35 (97.1%) | 12.14 (1.46 – 100.7) | 0.02 | 15.16 (1.31 – 175.06) | 0.03 |
| South Asia, East Asia & Pacific | 71 / 76 (93.4%) | 5.07 (1.59 – 16.16) | 0.006 | 15.43 (2.38 – 100.00) | 0.004 |
| **Years since arrival** | - | 1.01 (0.97 – 1.06) | 0.63 | 0.91 (0.83 – 1.00) | 0.06 |
| **Previous measles vaccination or infection** |  |  |  |  |  |
| No / Don't know | 97 / 106 (91.5%) | - | - | - | - |
| Yes | 36 / 43 (83.7%) | 0.47 (0.17 – 1.38) | 0.17 | 0.54 (0.12 – 2.54) | 0.44 |
| **Migrant status** |  |  |  |  |  |
| Non-refugee / asylum seeker | 118 / 133 (88.7%) | - | - | - | - |
| Refugee / asylum seeker | 15 / 16 (93.8%) | 1.91 (0.23 – 15.48) | 0.55 | 2.42 (0.17 – 34.82) | 0.52 |

## Self-reported vaccination and disease history and concordance analysis

Over two-thirds of the population (rubella 70.5%, measles 69.1%, varicella 67.1%) were unsure of their vaccination status. Similar proportions were unsure whether they had any history of these diseases (rubella 66.4%, measles 61.7% and varicella 47.7%) (Table 3).

**Table 3.** Self-reported vaccination and disease history stratified by serology result.

| **Measles** |  |  |
| --- | --- | --- |
| **History of vaccination** | **IgG positive** | **IgG negative** |
| No | 13 (9.8%) | 0 (0.0%) |
| Yes | 27 (20.3%) | 6 (37.5%) |
| Don't know | 93 (69.9%) | 10 (62.5%) |
| **History of infection** |  |  |
| No | 35 (26.3%) | 4 (25.0%) |
| Yes | 13 (9.8%) | 5 (31.3%) |
| Don't know | 85 (63.9%) | 7 (43.8%) |
| **History of disease or infection\*** |  |  |
| No | 23 (17.3%) | 4 (25.0%) |
| Yes | 36 (27.1%) | 7 (43.8%) |
| Don't know | 74 (55.6%) | 5 (31.3%) |
| **Rubella** |  |  |
| **History of vaccination** | **IgG positive** | **IgG negative** |
| No | 12 (8.7%) | 1 (9.1%) |
| Yes | 29 (21.0%) | 2 (18.8%) |
| Don't know | 97 (70.3%) | 8 (72.7%) |
| **History of infection** |  |  |
| No | 31 (22.5%) | 4 (36.4%) |
| Yes | 14 (10.1%) | 1 (9.1%) |
| Don't know | 93 (67.4%) | 6 (54.6%) |
| **History of disease or infection\*** |  |  |
| No | 25 (18.1%) | 3 (27.3%) |
| Yes | 33 (23.9%) | 2 (18.2%) |
| Don't know | 80 (58.0%) | 6 (54.6%) |
| **Varicella** |  |  |
| **History of vaccination** | **IgG positive** | **IgG negative** |
| No | 18 (12.3%) | 0 (0.0%) |
| Yes | 31 (21.2%) | 0 (0.0%) |
| Don't know | 97 (66.4%) | 3 (100.0%) |
| **History of infection** |  |  |
| No | 23 (15.8%) | 0 (0.0%) |
| Yes | 54 (37.0%) | 1 (33.3%) |
| Don't know | 69 (47.3%) | 2 (66.7%) |
| **History of disease or infection\*** |  |  |
| No | 17 (11.6%) | 0 (0.0%) |
| Yes | 67 (45.9%) | 1 (33.3%) |
| Don't know | 62 (42.5%) | 2 (66.7%) |

\*Don't know = those answering don't know to both history of vaccination and history of infection

Concordance between self-reported vaccination and disease history and serostatus is shown in Table 4. There was no agreement between a combined measure of vaccination and disease with serostatus (K: measles -0.05, varicella 0.01, rubella 0.03). The same predictor had a PPV of 83.7% for measles, 98.5% for varicella and 94.3% for rubella. NPV was less than 10% for all diseases.

**Table 4.** Concordance of self-reported vaccination and infection history with serology results.

| **Measles** |  | **IgG Positive** | **IgG Negative** | **PPV % (95%CI)** | **NPV % (95% CI)** | **Cohen's kappa** |
| --- | --- | --- | --- | --- | --- | --- |
| Disease | Yes | 13 | 5 | 72.2% (46.5 – 90.3%) | 8.4% (4.3 – 14.5%) | -0.05 |
|  | No* | 120 | 11 |  |  |  |
| Vaccine | Yes | 27 | 6 | 81.8% (64.5 – 93.0%) | 8.6% (4.2 – 15.3%) | -0.04 |
|  | No* | 106 | 10 |  |  |  |
| Combined D/V† | Yes | 36 | 7 | 83.7% (69.3% - 93.2%) | 8.5% (4.0 – 15.5%) | -0.05 |
|  | No* | 97 | 9 |  |  |  |
| **VZV** |  | **IgG Positive** | **IgG Negative** | **PPV % (95%CI)** | **NPV % (95% CI)** |  |
| Disease | Yes | 54 | 1 | 98.2% (90.3 – 100.0%) | 2.1% (0.3 – 7.5%) | 0.00 |
|  | No* | 92 | 2 |  |  |  |
| Vaccine | Yes | 31 | 0 | 100.0% (88.8% - 100.0%) | 2.5% (0.5 – 7.3%) | 0.01 |
|  | No* | 115 | 3 |  |  |  |
| Combined D/V† | Yes | 67 | 1 | 98.5% (92.1 – 100.0%) | 2.5% (0.3 – 8.6%) | 0.01 |
|  | No* | 79 | 2 |  |  |  |
| **Rubella** |  | **IgG Positive** | **IgG Negative** | **PPV % (95%CI)** | **NPV % (95% CI)** |  |
| Disease | Yes | 14 | 1 | 93.3% (68.1 – 99.8%) | 7.5% (3.6 – 13.3%) | 0.00 |
|  | No* | 124 | 10 |  |  |  |
| Vaccine | Yes | 29 | 2 | 93.6% (78.6 – 99.2%) | 7.6% (3.6 – 14.0%) | 0.01 |
|  | No* | 109 | 9 |  |  |  |
| Combined D/V† | Yes | 33 | 2 | 94.3% (80.8 – 99.3%) | 7.9% (3.7 – 14.5%) | 0.03 |
|  | No* | 105 | 9 |  |  |  |

† Yes = those answering yes to history of vaccination OR disease, No = those answering no or don't know to history of vaccination AND disease. \* those answering no combined with those answering don't know. D/V = disease or vaccination. PPV = positive predictive value, NPV = negative predictive value.

A sensitivity analysis excluding those unsure of both their disease and vaccination status shows little change in significant findings (Supplementary Table 3).

## Discussion

Migrants in Europe may be an under-immunised group which could be on account of disruption of health systems or receipt of insufficient vaccine dosage in their countries of origin, limited access to catch-up vaccination services in host countries and vaccine hesitancy (2-3, 6-8). The problem is compounded by the absence of reliable immunisation records for many migrant groups, making decisions regarding catch-up vaccination difficult. Recent guidelines from the European Commission on Disease Prevention and Control (ECDC) (9) and Public Health England (PHE) (10) recommend catch-up vaccination for adult migrants with no or incomplete vaccination records. While these guidelines make clinical decision-making easier, concerns have been raised about the possibility of over-vaccinating already protected individuals (3).

The present study was therefore conducted to rapidly establish preliminary data on seroprotection against measles, rubella and varicella amongst adult migrants living in the UK. We found that seroprotection rate for measles was below the HIT and that migrants from Europe and Central Asia (the majority of whom were from Eastern Europe) were least likely to have seroprotection against measles is in-line with previous European serosurveys (11, 12). Explanations for this observation may be found in the disturbance of health services by civil wars and low uptake of immunisation due to vaccine hesitancy (11, 13). Our findings would support targeted catch-up vaccination to adult migrants, for example those coming from endemic regions/countries within Europe. Additionally, our results highlight the concerns around vaccine hesitancy that may affect uptake of the SARS-CoV-2 vaccine. Widespread distribution of this vaccine, including in hard-to-reach groups such as migrants will be critical to slowing the spread of COVID-19. Targeted public health messaging aimed at particular migrant groups may provide an effective method of improving both routine and SARS-CoV-2 vaccine coverage.

We found increasing age was associated with increased likelihood of seroprotection against VPDs. An explanation for this could be decreased persistence of vaccine induced seroprotection compared to that induced by natural infection (14).

We assessed the value of self-reported vaccination history in predicting VPD serostatus. PPV was over 80% for all studied VPDs, largely due to the low number of participants that reported a history of vaccination or disease but did not have serological evidence of either event which, in turn, is due to the high seroprotection rate for all diseases. Additionally, NPVs were low and kappa coefficients showed no agreement between self-reported histories and serostatus, indicating the unreliability of self-reporting for ascertaining serostatus. Future studies should focus on comparing the cost effectiveness of checking serostatus and vaccinating the unprotected with offering catch-up vaccination to all migrants with an unclear vaccination record (9, 10).

Further, larger studies examining methods of enhancing vaccine coverage (e.g. incentivising catch-up vaccination in primary care) along with qualitative studies addressing issues such as vaccine hesitancy are urgently required. Attending to these problems is critical in light of the COVID-19 pandemic, where mass vaccination programmes (inclusive of hard-to-reach migrant groups), will be paramount in slowing transmission of infection.

Our study has limitations, including our small sample size. For the purposes of regression and concordance analyses we amalgamated those answering “no” and those answering “don’t know” to questions concerning vaccine/disease history. Given that a large proportion of migrants were unsure of their vaccine/disease status, excluding these individuals would have significantly reduced the number of observations in each analysis. To ensure this did not have a substantial impact on our findings we conducted a sensitivity analysis excluding those who were unsure of their status.

In conclusion, seroprotection rate for measles was below the HIT in UK migrants and younger migrants from Europe and Central Asia may be at high risk of contracting VPDs. There was no agreement between self-reported vaccination and disease history and serostatus. These findings should be used to inform targeted approaches to offering catch-up vaccination, taking into account demographic risk profile. Such an approach may provide an effective method of improving vaccine coverage in high risk migrant populations and warrants urgent investigation.

## Data Availability

Data requests can be made to the Chief Investigator and will be considered in conjunction with the Sponsor and Caldicott guardian.

## Author contributions

MP conceived the idea for the study with input into the protocol from SH. MG, CAM, MJW, KE, VR and AJL collected the data. CAM analysed the data. MG and CAM drafted the manuscript. All authors contributed to study planning and management, and revision of the manuscript and were in agreement to submit it for publication.

