## Supplementary Information for "Risk of vaccine preventable diseases in UK migrants: a serosurvey and concordance analysis, 2020"

**Supplementary table 1: Participants’ countries of birth**

| **Region of Origin** (in manuscript) | **Country of Birth** | **n (%)** |
| --- | --- | --- |
| **Europe & Central Asia**  (n=38) | Albania  Bulgaria  Germany  Ireland  Italy  Moldova  Poland  Uzbekistan | 1 (2.6%)  3 (7.9%)  1 (2.6%)  4 (10.5%)  1 (2.6%)  1 (2.6%)  26 (68.4%)  1 (2.6%) |
| **Africa & Middle East**  (n=35) | Democratic Republic of Congo  Ethiopia  Gambia  Iran  Iraq  Kenya  Libya  Malawi  Morocco  Mozambique  Nigeria  Somalia  South Sudan  Tanzania  Uganda  Zanzibar  Zimbabwe | 1 (2.9%)  2 (5.7%)  1 (2.9%)  2 (5.7%)  4 (11.4%)  5 (14.3%)  1 (2.9%)  2 (5.7%)  3 (8.5%)  1 (2.9%)  3 (8.6%)  1 (2.9%)  1 (2.9%)  2 (5.7%)  1 (2.9%)  1 (2.9%)  4 (11.4%) |
| **South Asia, East Asia & Pacific**  (n=76) | Afghanistan  Bangladesh  Burma  China  Fiji  Hong Kong  India  Pakistan  Sri Lanka  Thailand | 3 (4.0%)  18 (23.7%)  1 (1.3%)  1 (1.3%)  1 (1.3%)  1 (1.3%)  38 (50.0%)  9 (11.8%)  3 (4.0%)  1 (1.3%) |

**Supplementary Table 2:** **Predictors of seroprotection against vaccine preventable diseases**

| **Variable** | **n seroprotected / total (%)**  **133 / 149 (89.3%)** | **Unadjusted OR (95% CI)** | **p value** | **Adjusted OR (95% CI)** | **p value** |
| --- | --- | --- | --- | --- | --- |
| **Age** | - | 1.04 (1.00 – 1.08) | 0.03 | 1.08 (1.02 – 1.14) | 0.005 |
| **Gender**  Male  Female | 40 / 48 (83.3%)  82 / 101 (81.2%) | -  0.86 (0.35 – 2.14) | -  0.75 | -  0.95 (0.36 – 2.55) | -  0.93 |
| **Region of origin**  Europe & Central Asia  Africa & Middle East  Asia & Pacific | 28 / 38 (73.7%)  32 / 35 (91.4%)  62 / 76 (81.5%) | -  3.81 (0.95 – 15.24)  1.58 (0.63 – 3.99) | -  0.06  0.33 | -  3.94 (0.79 – 19.69)  1.57 (0.46 – 5.32) | -  0.10  0.47 |
| **Years since arrival** | - | 1.01 (0.97 – 1.05) | 0.64 | 0.96 (0.91 – 1.01) | 0.12 |
| **Previous vaccination or infection**  **Measles**  No / Don’t know  Yes  **Varicella**  No / Don’t know  Yes  **Rubella**  No / Don’t know  Yes | 89 / 106 (84.0%)  33 / 43 (76.7%)  71 / 81 (87.7%)  51 / 68 (75.0%)  96 / 114 (84.2%)  26 / 35 (74.3%) | -  0.63 (0.26 – 1.51)  -  0.42 (0.18 – 1.00)  -  0.54 (0.22 – 1.35) | -  0.30  -  0.05  -  0.19 | -  0.61 (0.14 – 2.56)  -  0.40 (0.14 – 1.17)  -  1.44 (0.30 – 6.98) | -  0.50  -  0.09  -  0.65 |

**Supplementary Table 3: Concordance of self-reported vaccination and infection history with serology results if those unsure of their status are excluded**

| **Measles** | | **IgG Positive** | **IgG Negative** | **PPV % (95%CI)** | **NPV % (95% CI)** | **Cohen’s kappa** |
| --- | --- | --- | --- | --- | --- | --- |
| Disease | Yes | 13 | 5 | 72.2% (46.5 – 90.3%) | 10.3% (2.9 – 24.2%) | -0.12 |
|  | No | 35 | 4 |  |  |  |
| Vaccine | Yes | 27 | 6 | 81.8% (64.5 – 93.0%) | 0.0% (0.0 – 24.7%) | -0.22 |
|  | No | 13 | 0 |  |  |  |
| Combined D/V^†^ | Yes | 36 | 7 | 83.7% (69.3% - 93.2%) | 0.0% (0.0 – 41.0%) | -0.16 |
|  | No | 7 | 0 |  |  |  |
| **VZV** | | **IgG Positive** | **IgG Negative** | **PPV % (95%CI)** | **NPV % (95% CI)** |  |
| Disease | Yes | 54 | 1 | 98.2% (90.3 – 100.0%) | 0.0% (0.0 – 100.0%) | -0.03 |
|  | No | 23 | 0 |  |  |  |
| Vaccine | Yes | 31 | 0 | 100.0% (88.8% - 100.0%) | 0.0% (0.0 – 18.5%) | 0.00 |
|  | No | 18 | 0 |  |  |  |
| Combined D/V^†^ | Yes | 67 | 1 | 98.5% (92.1 – 100.0%) | 0.0% (0.0 – 33.6%) | -0.02 |
|  | No | 9 | 0 |  |  |  |
| **Rubella** | | **IgG Positive** | **IgG Negative** | **PPV % (95%CI)** | **NPV % (95% CI)** |  |
| Disease | Yes | 14 | 1 | 93.3% (68.1 – 99.8%) | 11.4% (3.2 – 26.7%) | 0.03 |
|  | No | 31 | 4 |  |  |  |
| Vaccine | Yes | 29 | 2 | 93.6% (78.6 – 99.2%) | 7.7% (0.2 – 36.0%) | 0.02 |
|  | No | 12 | 1 |  |  |  |
| Combined D/V^†^ | Yes | 33 | 2 | 94.3% (80.8 – 99.3%) | 14.3% (0.4 – 57.9%) | 0.11 |
|  | No | 6 | 1 |  |  |  |

† Yes = those answering yes to history of vaccination OR disease, No = those answering no to history of vaccination AND disease. D/V = disease or vaccination. PPV = positive predictive value, NPV = negative predictive value

**Supplementary text**

**Recruitment sites**

Recruitment took place at four different sites: the Department of Infection and HIV Medicine, University Hospitals Leicester NHS Trust, Leicester City of Sanctuary walk-in centre (which provides support to asylum seekers and refugees), Leicester College and the St. Paul’s Polish Roman Catholic Church.

**Laboratory methods**

Measles and Varicella IgG were analysed using the Liaison XL assays, while the Rubella IgG was tested on the ADVIA Centaur XP according to the manufacturer’s instructions.
